# Are the Items of the Starkstein Apathy Scale Fit for the Purpose of Measuring Apathy Post-Stroke?

**DOI:** 10.1101/2021.05.05.21256484

**Authors:** Stanley Hum, Lesley K. Fellows, Christiane Lourenco, Nancy E. Mayo

## Abstract

Given the importance of apathy for stroke, we felt it was time to scrutinize the commonly used Starkstein Apathy Scale (SAS) for psychometric evidence that it is fit for this purpose. The objectives were to: (i) estimate the extent to which the SAS items fit a hierarchical continuum of the Rasch Model; and (ii) estimate the strength of the relationships between the Rasch analysed SAS and converging constructs related to stroke outcomes.

**Methods:** Data on 238 people with stroke (mean age=63.1 years (SD=12.1) women=37.4%) from a clinical trial of a community-based intervention targeting participation were available at 5 time points yielding 856 SAS questionnaires. SAS has 14 items, rated on a 4-point scale with higher values indicating more apathy. Psychometric properties were tested using Rasch partial-credit model, correlation, and regression. The construct was modeled as motivation with items rescored as high is better.

**Results:** Rasch analysis indicated that the response options were disordered for 8/14 items, pointing to unreliability in the interpretation of the response options; they were consequently reduced from 4 to 3. Only 9/14 items fit the Rasch model and therefore suitable for creating a total score. The new rSAS was deemed unidimensional (residual correlations: < 0.3), reasonably reliable (person separation index: 0.74), with item-locations uniform across time, age, sex, and education. However, 30% of scores were >2 SD above the standardized mean but only 2/9 items covered this range (construct mistargeting).

Apathy (rSAS/SAS) was correlated weakly with anxiety/depression and uncorrelated with physical capacity. Regression showed that the effect of apathy on participation and health perception was similar for rSAS/SAS versions: R^2^ participation measures ranged from 0.11 to 0.29; R^2^ for health perception was ∼0.25.

When placed on the same scale (0-42), rSAS value was 6.5 units lower than SAS value with minimal floor/ceiling effects. Estimated change over time was identical (0.12 units/month) which was not substantial (1.44 units/year) but greater than expected assuming no change (t: 3.6 and 2.4).

**Conclusion:** The retained items of the rSAS targeted behaviours more than beliefs and results support the rSAS as a robust measure of apathy in people with chronic stroke.

## Introduction

Apathy is a defining feature in many common neurological conditions, including Parkinson’s Disease, Alzheimer’s Disease, and stroke (1, 2) A meta-analysis of 24 studies found that apathy occurred in 30 to 40% of stroke patients(3). Mayo et al. estimated from an inception cohort that 20% of patients had apathy, as reported by a close companion, at some point in the first year post-stroke. They also found that apathy strongly affected recovery(4). Apathy has also been shown to affect health related quality of life (HRQL) in patients with stroke(5).

Although apathy is recognized as common and clinically important, there have been challenges in defining and measuring this construct. Marin originally described apathy as the “lack of motivation not attributable to disturbance of intellect, emotion, or level of consciousness”(6) and operationalized the definition as “a state characterized by simultaneous diminution in the overt behavioral, cognitive, and emotional concomitants of goal-directed behavior”(7). Medical diagnostic criteria were developed for apathy in 2009 and revised in 2018. The 2018 consensus group largely echoed Marin’s description, defining apathy as a quantitative reduction of goal-directed activity either in behavioral, cognitive, emotional, or social dimensions in comparison to the patient’s previous level of functioning in these areas. They also indicated that these changes may be reported by patients themselves or be based on the observations of others.

Practically, apathy can be considered as a diagnosis (i.e. a binary outcome) and as a state that can be measured along a continuum. Measures that include a series of questions (items) or structured interview components about apathy behaviours, are typically used for both diagnosis and measurement.

Such questionnaires assess apathy from the perspectives of: (i) the patient using a patient-reported outcome (PRO) or a self-reported outcome (SRO); (ii) an observer who is usually a significant other (ObsRO); or (iii) a health care professional using a clinician reported outcome (ClinRO)(8). There is an important distinction between PRO and SRO. The answer to the questions in a PRO can only be provided by the person without interpretation from anyone else(9). In SRO, the response given by the person can be amended based on other information that may not have been provided by the patient(8).

A continuous measurement scale is constructed by assigning numerical labels to the ordinal item-responses (e.g. not at all, slightly, some, and a lot) and summing these labels, assuming they have mathematical properties, which they may not. The continuous scale can be used to quantify severity and change over time. A specific cut-point on this continuum is used for diagnostic classification.

Notwithstanding the consensus on defining apathy, there is a plethora of apathy questionnaires and no gold standard(10). For example, a recent systematic review by Carrozzino 2019(11) identified 13 different apathy measures used in Parkinson’s Disease research. Most of these have also been used in other neurological conditions, including stroke, without the due diligence required to ensure that the interpretation of the values apply to these other populations(11).

As for many neuropsychiatric constructs, existing questionnaires were generally developed based on expert clinical input without a strong emphasis on psychometrics or patient experience to inform content. Classical approaches to psychometrics were applied after the items and the response options were set. The focus was on statistical homogeneity of the items (internal consistency expressed by Cronbach’s alpha) and factor structure. Dimensionality of these measures was felt necessary to cover the construct, but not always reflected in the scoring. Further the psychometrics underlying the total score were not scrutinized; measures were formed as a simple sum of the ordinally-labeled response options. These approaches have well-known drawbacks: A high internal consistency can arise from redundant items, and change on one item will yield similar changes in the redundant items resulting in an overestimate of the calculated amount of change. These legacy measures are now being scrutinized in light of modern psychometric methods such as Item Response Theory (IRT) and Rasch Measurement Theory (RMT)(12, 13).

RMT estimates the extent to which a set of items fits an underlying linear hierarchy (Rasch Model); items that do not fit this hypothesized model should not be included in the total score until revised(14). Thus, psychometrics come into play before the items and response options are set. RMT tests whether response options are functioning as expected in terms of representing more (or less) of the construct and if not, modifications are made. Rasch analysis is also used to transform the ordinal scores of the response options to have mathematical interval-like properties, rather than just numerical labels, allowing a legitimate total score to be derived. This approach also allows testing whether the items reflect more than one dimension and, therefore, whether the construct is best reflected by multiple measures.

Another key feature of these modern psychometric methods is the ability to test whether the items have the same mathematical interpretation in different subpopulations, such as those defined by sex or gender, language, or by different disease groups(15, 16). The additional information on how the items respond is useful in creating a measure with strong properties, needed particularly for evaluating change over time.

The most commonly used self-reported questionnaire for assessing apathy in stroke are the 18 item Apathy Evaluation Scale (AES) developed by Marin(7) in 1991 and the Starkstein Apathy Scale (SAS)(17) which is based on a preliminary version of the AES. The SAS consists of a combination of 14 (PRO/SRO) items with a four-point ordinal scale (0=not at all; 1=slightly; 2=some; and 3=a lot), with a higher summed total score indicating more apathy. We were unable to find any studies of the psychometric properties of the SAS in stroke, although it has been used in that population(18). Given the importance of apathy for stroke, we felt it was time to scrutinize and, if needed, improve the SAS using modern psychometric approaches. The global aim of this study was to contribute evidence supporting the use of the SAS in people with stroke. The specific objectives were to: (i) to estimate the extent to which the items of the SAS fit a hierarchical continuum based on the Rasch Model; and (ii) to estimate the strength of the relationship between the Rasch analysed SAS and converging constructs related to stroke outcomes.

## Methods

This study is a secondary analysis of an existing dataset from a clinical trial assessing a community-based, structured, program targeting participation in individuals post-stroke(19). A sample of 238 English speaking participants living in the community having experienced a stroke within five years responded to the SAS at study entry (baseline=0 month) and up to 4 more times (at 3, 6, 12, and > 12 months). A total of 856 SAS questionnaires were completed by the cohort over these time points.

### Measurement

The World Health Organization’s (WHO) international classification for functioning, disability, and health (ICF) model) was used as the measurement framework and to structure the analyses as shown in Figure 1. Apathy is not part of the ICF, but motivation is listed as an impairment (b1301: mental functions that produce the incentive to act; the conscious or unconscious driving force for action). Apathy is part of the WHO international classification of diseases (ICD-10), listed as a diagnostic category not as a function (R45 symptoms and signs involving emotional state, specifically R45.3, demoralization and apathy). Apathy was represented by the original SAS with 14 items measured on a 4-point severity scale.

**Figure 1.**
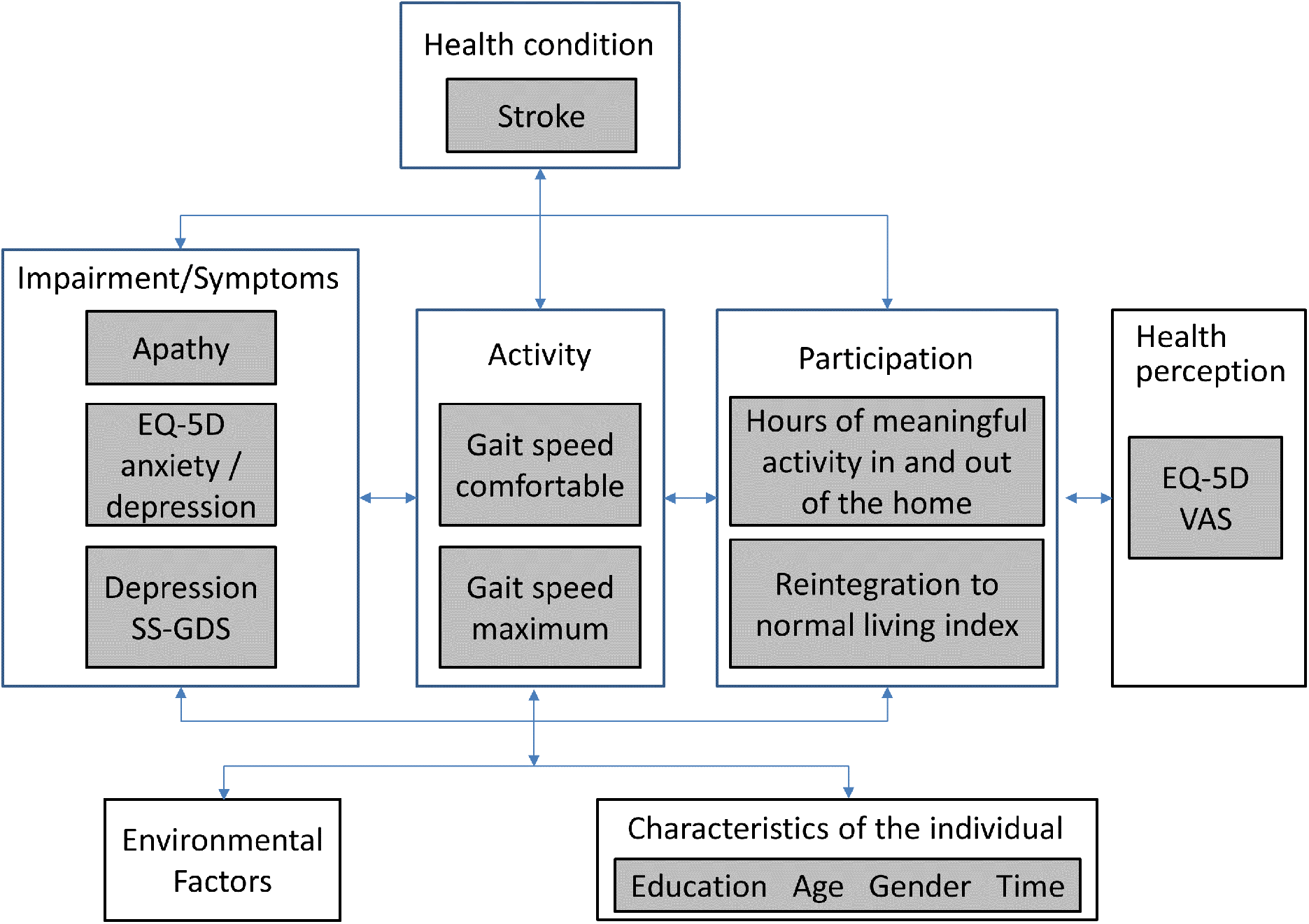
WHO International Classification of Functioning, Disability and Health (ICF) for the measurement framework

The literature supports a relationship between apathy and depression (both diagnostic categories; depression is ICD-10 F32-F34). Here, we included two measures of depressive symptoms (ICF b1265: impairment of optimism as defined by mental functions that produce a personal disposition that is cheerful, buoyant and hopeful, as contrasted to being downhearted, gloomy and despairing): Stroke-Specific Geriatric Depression Scale (SS-GDS)(20) and the anxiety/depression item of the Euroqol EQ-5D(21).

There is a very strong effect of impairment of apathy/motivation on physical function, participation and self-rated health(22). Physical function was assessed here with measured gait speed (comfortable and maximum), hypothesized to be less affected by motivation as it is a measure of capacity to walk a short distance (5 meters). Two measures of participation were available: Community Healthy Activities Model Program for Seniors [CHAMPS](23) using hours of meaningful activity in and outside the home as the metric and the Reintegration to Normal Living Index [RNL](24). The EQ-5D VAS (visual analog scale: 0 - death to 100 - perfect health) was the measure of health perception. Personal factors under consideration were gender, time (0; 3; 6; 12; > 12 months), age (<50; 50-60; 61-70; 71-80; >80 years), and education (≤ 12 years; > 12 years).

## Statistical methods

Rasch analysis was used to test whether the items on the SAS fit the Rasch model(25). The steps and requirements for Rasch analysis were adopted from published guidelines(15, 16) and are detailed in Figure 2/2a. RUMM2020 version 4.1 software was used to fit a partial credit model. The process is iterative and items that do not fit are removed one at a time until all items fit the model.

**Figure 2.**
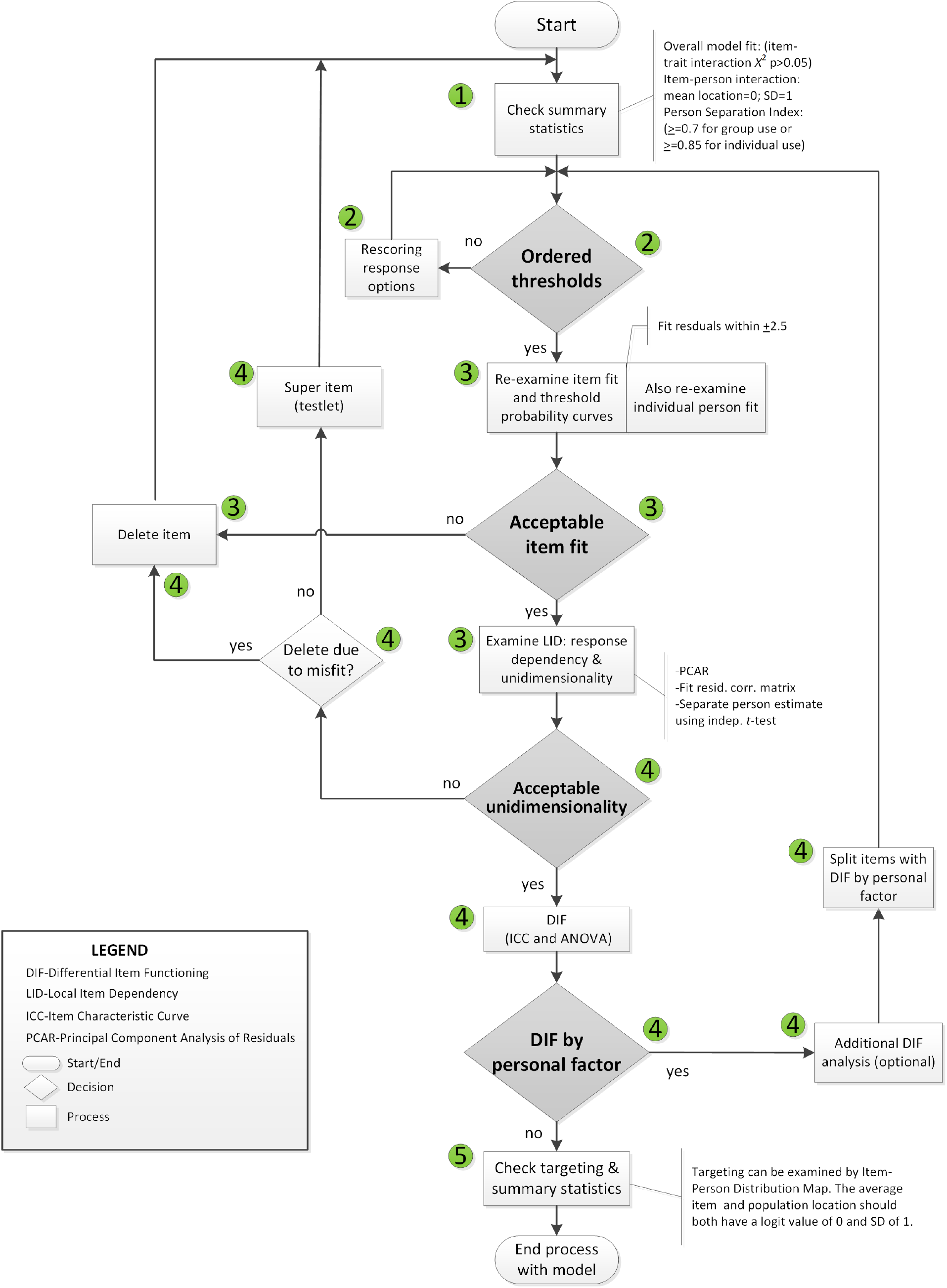
Rasch Analysis Decision Flowchart

**Figure 2a.**
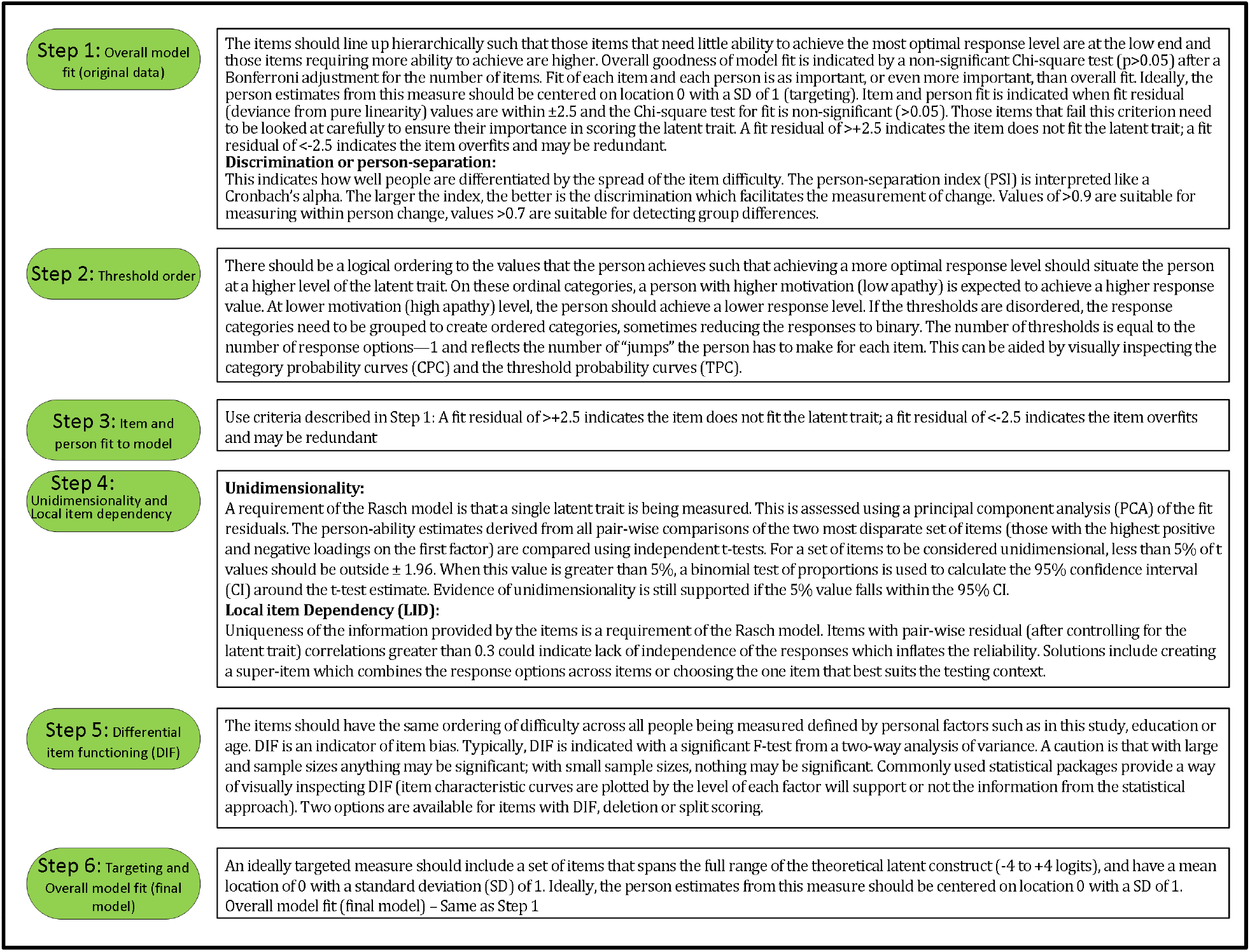
Explanation of steps taken to fit the data to the Rasch model.

Rasch analysis was conducted on all 14 items, initially. The number of observations available for Rasch analysis was 856 arising from 238 people with stroke, more than adequate to estimate item and person difficulties with a high degree of confidence level (99%) within a precision of ± 0.5 logits(26).

The lack of independence in the observations owing to repeated measures does not affect item locations on the hierarchy. In fact, it is an advantage in that the effect of repeated measures (time) can be tested using differential item functioning (DIF), where the hypothesis is the ordering of the items is unaffected by time. This is a valuable psychometric property for the estimation of change and is used to distinguish between change from response shift and true change and also to identify if there is a learning effect(27).

Correlation coefficients were calculated between the revised SAS total score (rSAS) derived from Rasch analysis for those constructs at the same ICF level, i.e. measures of depressive symptoms (polyserial correlation for ED-5D item anxiety/depression and Pearson correlation for SS-GDS). As these measures are theorized to relate to the same latent construct, the following criteria were used to qualify the strength: strong ≥0.8; moderate 0.50-0.80; weak <0.50.(28). For downstream outcomes (participation and health perception), linear regression was used with adjustment for age, sex and gait speed. Here the interpretation of the strength of correlation coefficients for novel relationships was used (strong: ≥0.5)

Linear regression was also used to estimate the extent to which the response of the stroke participants to the community-based participation-targeted intervention (from the original study) differed when measured using SAS and rSAS. The regression parameter for slope (β) quantified linear growth over time and the *t*-statistic, derived from the ratio of the β to its standard error (SE) was used for effect size. As regression models used explanatory variables with different measurement scales, to facilitate comparison standardized regression coefficients were used.

Generalized estimating equations (GEE) was used to make a direct comparison of change over time between the two measures. Here the value for apathy was the outcome and the explanatory variables were the version of the measure used to derive the score (original SAS and rSAS), time, and age and sex as adjustment variables. GEE considers the clustering of apathy value in the measure and adjusts the error variance accordingly.

## Results

Participant characteristics of the sample are summarized in Table I. On average, participants had a stroke 2.4 years before study entry. Table II summarizes the results of the Rasch analysis. Disordered thresholds were observed in 8 of the 14 items. Disordered thresholds were consistently observed for “slightly” and “some” requiring the response options to be collapsed. Only item #8 retained the original response options. The remaining items were rescored due to poor spacing between Threshold Probability Curves (TPC) or poor fit between the observed values and the expected TPC. Iterations of the Rasch analysis are summarized in Table III. After rescoring, the 14 items SAS still did not fit the Rasch model. The overall chi-square value for item-trait interaction was statistically significant [X^2^(70)=510.767 p value<0.000](Table III). Item #3 and #5 had initial fit residuals of +10.8 and +5.9 respectively and were consistently > +2.5 even after iterative adjustments with respect to the model and were deleted because of lack of fit to the hierarchical construct. Items #13 and #6 were iteratively deleted for the same reason. Item #11 (Are you unconcerned with many things?) did fit the Rasch model but had similar location (difficulty) values as item #10 (Are you indifferent to things?) with < 0.2 logit difference between the items and was deleted because it was poorly worded.

**Table I.**
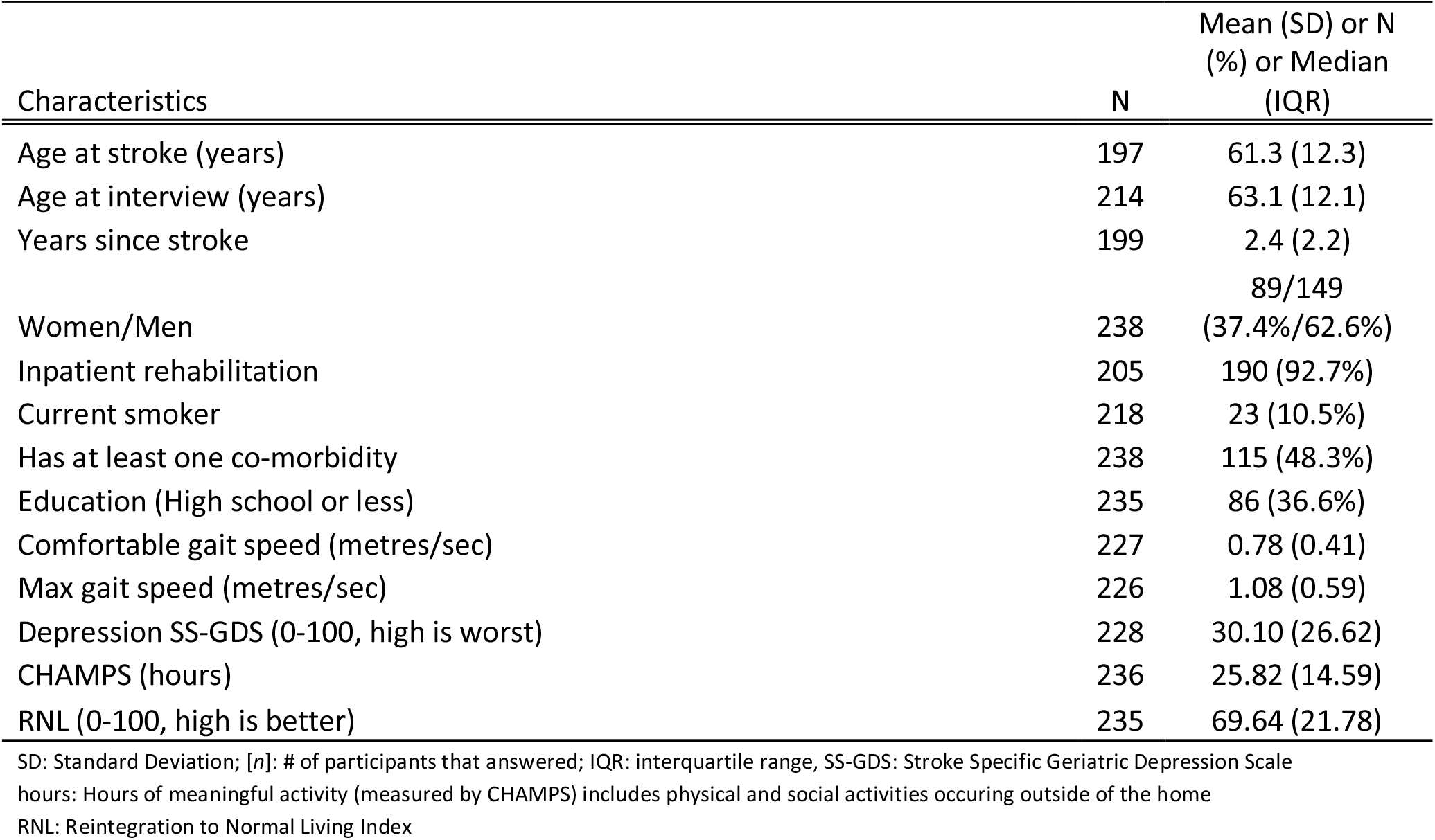
Demographic and clinical characteristics of the sample (n=238)

| Characteristics | N | Mean (SD) or N (%) or Median (IQR) |
| --- | --- | --- |
| Age at stroke (years) | 197 | 61.3 (12.3) |
| Age at interview (years) | 214 | 63.1 (12.1) |
| Years since stroke | 199 | 2.4 (2.2) |
| Women/Men | 238 | 89/149 (37.4%/62.6%) |
| Inpatient rehabilitation | 205 | 190 (92.7%) |
| Current smoker | 218 | 23 (10.5%) |
| Has at least one co-morbidity | 238 | 115 (48.3%) |
| Education (High school or less) | 235 | 86 (36.6%) |
| Comfortable gait speed (metres/sec) | 227 | 0.78 (0.41) |
| Max gait speed (metres/sec) | 226 | 1.08 (0.59) |
| Depression SS-GDS (0-100, high is worst) | 228 | 30.10 (26.62) |
| CHAMPS (hours) | 236 | 25.82 (14.59) |
| RNL (0-100, high is better) | 235 | 69.64 (21.78) |

SD: Standard Deviation; [n]: # of participants that answered; IQR: interquartile range, SS-GDS: Stroke Specific Geriatric Depression Scale
hours: Hours of meaningful activity (measured by CHAMPS) includes physical and social activities occurring outside of the home
RNL: Reintegration to Normal Living Index

**Table II.** Starkstein’s Apathy Scale items retained/deleted

| Item | Description | response options | Location | SE | Fit Residual |
| --- | --- | --- | --- | --- | --- |
| I0001 | Are you interested in learning new things? | 0-1-1-2 | -0.088 | 0.071 | 0.642 |
| *I0002 | Does anything interest you? | 0-1-1-2 | -0.065 | 0.070 | -1.391 |
| *I0003 | 3. Are you concerned about your condition? | 0-1-1-2 | deleted-construct misfit |  |  |
| I0004 | Do you put much effort into things? | 0-1-1-2 | -0.179 | 0.072 | 0.037 |
| *I0005 | 5. Are you always looking for something to do? | 0-1-1-2 | deleted-construct misfit |  |  |
| *I0006 | 6. Do you have plans and goals for the future? | 0-1-1-2 | deleted-construct misfit |  |  |
| I0007 | 7. Do you have motivation? | 0-1-1-2 | 0.514 | 0.069 | -2.482 |
| I0008 | Do you have the energy for daily activities? | 0-1-2-3 | 0.740 | 0.053 | 0.805 |
| *I0009 | Does someone have to tell you what to do each day? | 0-1-1-2 | -0.596 | 0.073 | 2.017 |
| I0010 | Are you indifferent to things? | 0-1-1-2 | -0.180 | 0.070 | 0.305 |
| I0011 | 11. Are you unconcerned with many things? | 0-1-1-2 | location redundancy with #10, also poorly worded |  |  |
| *I0012 | Do you need a push to get started on things? | 0-1-1-2 | 0.277 | 0.067 | -0.174 |
| *I0013 | 13. Are you neither happy nor sad, just in between? | 0-1-1-2 | deleted-construct misfit |  |  |
| *I0014 | 14. Would you consider yourself apathetic? | 0-1-1-2 | -0.423 | 0.070 | -2.454 |

\*Indicates items with disordered threshold requiring rescoring response options

**Table III.** Summary fit statistics for Starkstein’s Apathy Scale

| Model | sample<br>size (n) | Overall model fit | Item<br>Location | Item fit<br>residual<br>Mean<br>(SD) | Person<br>Location | Person fit<br>residual<br>Mean (SD) | PSI |
| --- | --- | --- | --- | --- | --- | --- | --- |
| 14<br>items | 856 | $\chi^2=510.767$, df=70,<br>p=0.0000 | 0.0<br>(0.461) | -0.871<br>(3.635) | 0.820<br>(0.695) | -0.166<br>(1.201) | 0.74 |
| 14<br>items | 856 | $\chi^2=422.249$, df=70,<br>p=0.0000 | 0.0<br>(0.678) | -0.003<br>(3.440) | 1.124<br>(0.946) | -0.284<br>(1.263) | 0.73 |
| 9 items | 809 | $\chi^2=89.103$, df=45,<br>p=0.000099 | 0.0<br>(0.433) | -0.299<br>(1.523) | 1.602<br>(1.274) | -0.333<br>(1.203) | 0.74 |

The remaining nine items met the requirements of the Rasch model with fit residuals between ±2.5 as shown in Table II. During iterative analysis, Item #7 and #14 had fit residuals < −2.5 indicating items over-discriminated the response pattern and may be redundant but in the final model the values were just within the cut-off (∼-2.4).

Local dependency was investigated using the correlation matrix of the residuals to examine response dependencies between items. All item-pair correlations were less than 0.3. A single construct, “apathy” was also supported with less than 5% of *t*-tests being significant. Reliability based on the person separation index (PSI) is 0.74.

Targeting was assessed using the person-item threshold distribution map for the nine SAS items as shown in Figure 3. The figure shows item thresholds were reasonably well distributed over ∼5 logits with a near normal distribution (logit range ∼-2.0 to 2.8). The sample population had a logit mean of 1.60; SD:1.3 and a distribution range between ∼-1.6 to 4.4 logits showing some individuals at the low apathy end of the scale were not measured as reliably as there were no items that extended into that range. As shown in Figure 3, ∼13% of respondents’ scores were higher than any item available for the assessment. Also, 30% of scores were >2 SD above the standardized mean but only 2/9 items covered this range.

**Figure 3.**
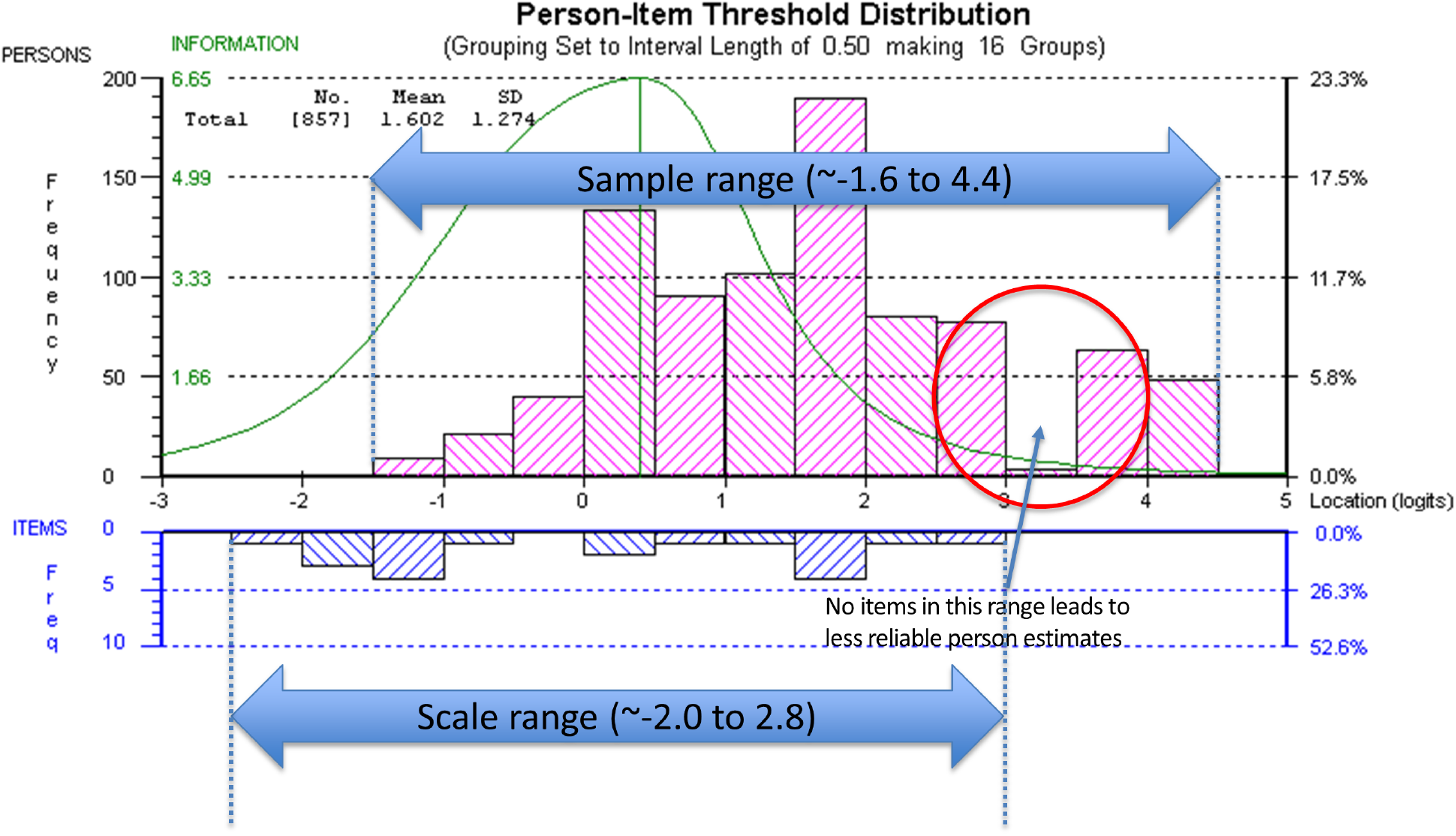
Person-Item Threshold Distribution Map

The summary statistics based on the final model with 9 items had an overall chi-square value for item-trait interaction that was still statistically significant [X^2^(45)=89.103 p value<0.0001](Table III).

As there are a large number of observations (n=809) making the analysis overly sensitive to the detection of even trivial misfit, five random samples of 300 each (with replacement) were drawn(29) and the average p-value for fit was non-significant (0.28), indicating global fit (Table Ia in the appendix). There was no substantial DIF by time, age, sex, or education in the 5 random samples.

The correlations between the original SAS or rSAS apathy total score with measures used to support interpretability are shown in Table IV. The strongest correlations (range −0.37 to −0.41) were observed between the convergent construct of depressed mood, measured using the EQ-5D anxiety/depression item and the SS-GDS. Neither apathy scale version was correlated with comfortable or maximum gait speed (m/s) with estimates ranging from 0.07 to 0.13.

**Table IV.** Relationship between rSAS apathy total score and measures used to support interpretability. Correlation of the original SAS and rSAS score with known outcomes to estimate convergent validity.

| variable name | N |  | Original<br>SAS score | p-value | rSAS<br>(9 items) | p-value |
| --- | --- | --- | --- | --- | --- | --- |
|  |  |  | correlation |  | correlation |  |
| anxiety/depression item EQ-5D | 238 | polyserial | -0.41 | <0.000 | -0.40 | <0.000 |
| SS-GDS (ladder 0-100) | 228 | Pearson | -0.37 | <0.000 | -0.40 | <0.000 |
| comfortable gait speed (m/s) | 227 | Pearson | 0.09 | 0.154 | 0.13 | 0.050 |
| max gait speed (m/s) | 226 | Pearson | 0.07 | 0.311 | 0.11 | 0.104 |

Table V shows the extent to which the two different versions of the SAS (original and rSAS) explain downstream outcomes of participation and perceived health adjusted for age, sex, and gait speed. For CHAMPS-hours of meaningful activity, the effect of apathy was similar for the two versions (SAS β:0.32, t:4.85) and for (rSAS β:0.29, t:4.27). This similarity held for the other downstream outcomes. R^2^ participation measures for SAS and rSAS ranged from 0.11 to 0.29, equivalent to correlations ranging from 0.34 to 0.54 (considered moderate to strong); R^2^ for health perception was approximately 0.25 (correlation 0.5, strong).

**Table V.** Regression of apathy (original total score or Rasch apathy score [rSAS]) as a predictor of downstream outcomes (participation/HRQL)

|  |  |  |  |  |  |  |  |  |  |
| --- | --- | --- | --- | --- | --- | --- | --- | --- | --- |
| **Participation (CHAMPS hours)** N=208<br>r=0.368; R<sup>2</sup>=0.135 \| F Change=7.930<br>Sig. F Change=0.000 |  |  |  |  | r=0.338; R<sup>2</sup>=0.114 F Change=6.560<br>Sig. F Change=0.000 |  |  |  |  |
| | | $\beta$ | t | Sig. | | | $\beta$ | t | Sig. |
| Original apathy score (SAS) |  | 0.32 | 4.85 | 0.000 | Rasch apathy score (rSAS) |  | 0.29 | 4.27 | 0.000 |
| Age at interview |  | -0.03 | -0.03 | 0.643 | Age at interview |  | -0.03 | -0.44 | 0.658 |
| Men(1) vs Women(0) |  | 0.07 | 1.12 | 0.266 | Men(1) vs Women(0) |  | 0.07 | 1.05 | 0.293 |
| Comfortable gait speed(m/s) |  | 0.14 | 2.08 | 0.039 | Comfortable gait speed(m/s) |  | 0.13 | 1.93 | 0.054 |
| **Participation (RNL)** N=207<br>r=0.544; R<sup>2</sup>=0.296 \| F Change=17.478<br>Sig. F Change=0.000 |  |  |  |  | r=0.526; R<sup>2</sup>=0.277 \| F Change=19.351<br>Sig. F Change=0.000 |  |  |  |  |
| | Model | $\beta$ | t | Sig. | | Model | $\beta$ | t | Sig. |
| Original apathy score (SAS) |  | 0.34 | 5.67 | 0.000 | Rasch apathy score (rSAS) |  | 0.31 | 5.11 | 0.000 |
| Age at interview |  | 0.24 | 4.03 | 0.000 | Age at interview |  | 0.24 | 3.93 | 0.000 |
| Men(1) vs Women(0) |  | -0.09 | -1.56 | 0.121 | Men(1) vs Women(0) |  | -0.10 | -1.61 | 0.109 |
| Comfortable gait speed(m/s) |  | 0.31 | 5.14 | 0.000 | Comfortable gait speed(m/s) |  | 0.30 | 4.89 | 0.000 |
| **Health perception (EQ-5D VAS)** N=207<br>r=0.492; R<sup>2</sup>=0.242 \| F Change=16.086<br>Sig. F Change=0.000 |  |  |  |  | r=0.497; R<sup>2</sup>=0.247 \| F Change=16.573<br>Sig. F Change=0.000 |  |  |  |  |
| | | $\beta$ | t | Sig. | | | $\beta$ | t | Sig. |
| Original apathy score (SAS) |  | 0.39 | 6.37 | 0.000 | Rasch apathy score (rSAS) |  | 0.40 | 6.51 | 0.000 |
| Age at interview |  | 0.14 | 2.21 | 0.028 | Age at interview |  | 0.13 | 2.13 | 0.034 |
| Men(1) vs Women(0) |  | 0.02 | 0.36 | 0.719 | Men(1) vs Women(0) |  | 0.02 | 0.33 | 0.742 |
| Comfortable gait speed(m/s) |  | 0.21 | 3.38 | 0.001 | Comfortable gait speed(m/s) |  | 0.19 | 3.08 | 0.002 |

Table VI shows key measurement properties of the two versions. GEE showed the 9-item rSAS (rescaled to be out of 42) produced a value that was 6.5 units lower than the full 14-item version with more variability. Floor and ceiling effects were minimal for the two versions. Estimated change over time was identical (0.12 units per month) which was not substantial (1.44 units per year) but greater than expected assuming no change (t: 3.6 and 2.4).

**Table VI.** Floor and Ceiling Effects and Responsiveness of the original SAS and rSAS

| N=238 | SAS | rSAS |
| --- | --- | --- |
| Items | 14 | 9 |
| Scoring range | 0-42 | 0-19 (rescaled to 0-42) |
| Mean (SD) [baseline] | 28.1 (6.5) | 21.3 (9.2) |
| Adjusted difference* | -6.5 (SE:0.25; t:26.10) |  |
| CV | 0.23 | 0.43 |
| Floor / Ceiling: n (%) | 0/1 (0%/0.4%) | 0/11 (0%/4.6%) |
| Change over time ($\beta$) | 0.12 | 0.12 |
| Effect size ($\beta$ / SE) | 3.6 | 2.4 |

\*Adjusted difference (SE) estimated using GEE.
CV = coefficient of variation; $\beta$ = standardized beta; SD = standard deviation; SE = standard error

## Discussion

This study found that 9 of the original 14 items of the SAS fit a linear hierarchy (the Rasch model) suitable for measuring apathy in people with stroke. The results of the Rasch analysis on the original four-point ordinal scale showed that these thresholds were not used in a manner consistent with endorsing more positive response option with decreasing apathy (increasing motivation) (i.e., disordered thresholds). On 8 of the 14 items, participants consistently had difficulty differentiating reliably between the middle two response options “slightly” and “some”. The rationale of having more response options is to try to be more precise in the assessment, but the observed disordered thresholds indicate that participants could not reliably distinguish between some choices, so this design choice was counter-productive. Not taking disordered thresholds into account can provide a false sense of reliability and increase measurement error.

The original SAS items measure several aspects of apathy according to Pedersen et al. such as: 1) diminished motivation (#7 and #12); 2) behavioral (#4, #5, #8, and #9); 3) cognitive (#1, #2, #6, #11); 4) emotional (#10 and #13); and 5) insight (#3 and #14) (30). Our analysis showed four items (#3, #5, #6, and #13) did not fit the latent construct and so were removed from the rSAS. The poor fit of item #3 “Are you concerned about your condition?” has also been described in three other Parkinson’s studies as unreliable or ambiguous(30-32).

Morita and Kannari 2016 found that items #11 and #13 were not reflective of apathy in Parkinson’s population(32). Our analysis also showed that item #13 “Are you neither happy nor sad, just in between?” did not fit the apathy construct and so was deleted from the measure. On the other hand, although item #11 “Are you unconcerned with many things?” did fit our model, it was so poorly worded that the decision was made to delete it. Item #5 “Are you always looking for something to do?” and item #6 “Do you have plans and goals for the future?” did not fit the apathy construct in the stroke population, possibly because these are features of stroke rather than apathy.

Fit residuals for items #7 and #14 were just within the cut-off of −2.5, indicating marginal over-fit to the Rasch model. These were retained in the rSAS version. These two items are “reverse worded items” such that they might be considered essentially the same question with one positively worded and the other one negatively worded. Using words with opposite meaning has been discussed as a means to control for acquiescence and social desirability bias(33) but also may have the effect of artificially increasing the reliability of the SAS.

Of the 9 items fitting the Rasch model, 7 are classified as self-report items as they query observable behaviors where the response provided by the patient could be amended based on other information independent of the patient. The only two PRO items in the rSAS version are item #7 and #14 asking the person directly if they think they are motivated or apathetic, respectively.

Our results did not show a correlation between apathy and tests of physical capacity (comfortable or maximum gait speed). A systematic review did report an association between apathy and increased disability post-stroke in most studies, however the disability outcomes were mostly related to activities of daily living which require effort. The authors also noted that they could not perform a quantitative meta-analysis due to the amount of heterogeneity in the outcomes and analyses used among the studies(3).

The literature supports depression as being distinct from apathy, but with some degree of overlap(4, 10). Previous work also suggests that apathy can coexist with depression to varying degree in stroke populations specifically(4). A review of several different apathy measures show low correlation with depression(10). Our hypothesis was that depression constructs would correlate weakly with the rSAS. The weak correlation observed between rSAS and depression outcomes (SS-GDS and EQ-5D anxiety /depression) provided further evidence that apathy is a distinct construct from depression. The modest correlation (∼0.4) between SS-GDS and rSAS may still be due to the inclusion of overlapping items asking about loss of interest, doing new things, and energy level contained in both questionnaires. It would be prudent to select apathy and depression scales without items querying these common features to avoid misclassification, given that apathy and depression can coexist.

In our study, the amount of variance in participation measures explained by either the SAS or rSAS was small and very similar (range 0.114 to 0.296, Table V). However, these 2 versions explained approximately 25% of the variance in health perception. This indicates that apathy has more to do with how people feel than what they do. What they do, may be influenced by family activities.

Our result is largely consistent with the reported estimates in the literature showing some association between apathy and HRQL in autoimmune, inherited, and neurodegenerative disorders; however, apathy was used as an outcome or as an exposure variable in the analysis in the different studies (5, 34-36).

The distribution of the items of the SAS along the latent trait of apathy does not match the distribution of the values on the latent trait observed in sample. We have conceived of the apathy construct to range from apathy (low end of the scale) to motivation (high end of the scale) with the latent trait standardized to have a mean of 0 and a SD of 1. The mean location along the latent trait with all 14 items after rescoring due to disordered thresholds was 1.0 (SD 0.9); with the rSAS of 9-items the mean location was 1.6 (SD: 1.2). This means that people had more motivation than the items were able to measure suggesting that to adequately measure the full range of the construct other items would be needed. Until a stroke-specific measure of the apathy-motivation construct can be developed, researchers could use the nine rSAS items.

### Future directions

This study showed that a 9-item version of the original 14-item SAS could be used to assess apathy in chronic stroke patients without loss of content coverage but with gain in mathematical properties. The lack of targeting of the items to the range of motivation shown in this sample indicates additional items are needed. In addition, since the development of the original SAS, recommendations for developing PROs have been made(9). This process requires input from patients, caregivers, and clinicians and verification that all requirements for a robust, mathematically sound measure are met. Many disciplines are revisiting their legacy measures to assess the extent to which they measure up to these new standards and also adopting new measures that meet these new standards.

## Data Availability

Academic researchers interested in obtaining the data can make a request to the corresponding author. A research proposal would need to be submitted to our Research Ethics Board and a data sharing agreement would be required between the collaborating institutions.

## Appendix

**Table Ia.** Summary fit statistics for Starkstein’s Apathy Scale based on 5 random sample each with n=300

| Model | sample<br>size (n) | Overall model fit | Item<br>Location | Item fit<br>residual<br>Mean (SD) | Person<br>Location | Person fit<br>residual<br>Mean<br>(SD) | PSI |
| --- | --- | --- | --- | --- | --- | --- | --- |
| sample1 | 282 | $\chi^2=39.547$, df=45,<br>p=0.701 | 0.0<br>(0.457) | -0.137<br>(1.095) | 1.680<br>(1.279) | -0.293<br>(1.095) | 0.74 |
| sample2 | 283 | $\chi^2=56.644$, df=45,<br>p=0.114287 | 0.0<br>(0.495) | -0.148<br>(1.010) | 1.656<br>(1.298) | -0.356<br>(1.190) | 0.74 |
| sample3 | 284 | $\chi^2=58.835$, df=45,<br>p=0.080793 | 0.0<br>(0.453) | -0.122<br>(0.976) | 1.612<br>(1.225) | -0.315<br>(1.132) | 0.72 |
| sample4 | 287 | $\chi^2=47.047$, df=45,<br>p=0.388697 | 0.0<br>(0.412) | -0.172<br>(0.978) | 1.479<br>(1.316) | 0.303<br>(1.183) | 0.75 |
| sample5 | 282 | $\chi^2=56.119$, df=45,<br>p=0.123756 | 0.0<br>(0.443) | 0.033<br>(1.149) | 1.562<br>(1.198) | -0.328<br>(1.193) | 0.72 |
| average |  | p=0.28 |  |  |  |  | 0.73 |

